# Sex Differences in Mortality and Treatment Utilization Across Cardiogenic Shock Phenotypes: A National Cohort Study

**DOI:** 10.64898/2026.05.26.26354172

**Authors:** Arthur Alencar, Xuan Li, Abdulgafar Ibrahim, Arundhati Sawant, Michael Bashir, Venkata Varshitha Bandi, Karan Bhatt, Ahmad Jalil, Vijay Chennareddy

**Affiliations:** Department of Internal Medicine, Baptist Memorial Hospital-Golden Triangle, Columbus, Mississippi, USA; Department of Internal Medicine, Baptist Memorial Hospital-Jackson, Jackson, Mississippi, USA; Department of Internal Medicine, Baptist Memorial Hospital-North Mississippi, Oxford, Mississippi, USA; Department of Internal Medicine, Texas Tech University Health Sciences Center El Paso, El Paso, Texas, USA

**Keywords:** cardiogenic shock, sex differences, mechanical circulatory support, mortality, heart failure, acute myocardial infarction

## Abstract

**Background:** Cardiogenic shock (CS) is a heterogeneous syndrome with diverse etiologies, treatment pathways, and outcomes. Prior studies of sex differences in CS have largely focused on acute myocardial infarction-related CS or evaluated CS as a single entity. Whether sex-based differences in outcomes and treatment utilization vary across distinct CS phenotypes remains incompletely defined.

**Methods:** We performed a retrospective cohort study using the National Inpatient Sample, a nationally representative all-payer database of United States hospitalizations. Adult hospitalizations with CS were identified using ICD-10-CM code R57.0 and categorized into clinically relevant phenotypes, including acute myocardial infarction (AMI), heart failure (HF), arrhythmia-related shock, myocarditis/Takotsubo, valvular disease, and other etiologies. Survey-weighted analyses accounting for the complex sampling design were used for primary analyses. The primary outcome was in-hospital mortality. Secondary outcomes included use of mechanical circulatory support (MCS) and mechanical ventilation. Propensity score-matched analyses were performed as sensitivity analyses.

**Results:** Among 254,691 weighted CS hospitalizations, 158,747 (62.3%) occurred in men and 95,896 (37.7%) in women. In survey-weighted analyses, women had higher in-hospital mortality in AMI-related CS (36.1% versus 31.3%; OR, 1.24; 95% CI, 1.19-1.28), HF-related CS (30.5% versus 25.8%; OR, 1.27; 95% CI, 1.23-1.30), and arrhythmia-related CS (37.3% versus 31.6%; OR, 1.28; 95% CI, 1.20-1.38). Women were less likely to receive ECMO (2.4% versus 2.9%), IABP/Impella (13.1% versus 18.9%), or any MCS (14.6% versus 20.4%), but were more likely to receive mechanical ventilation (44.9% versus 42.9%). In propensity-matched analyses, mortality differences were attenuated but persisted in AMI-related, HF-related, and valvular CS.

**Conclusions:** Sex differences in CS outcomes and treatment utilization are strongly phenotype dependent. Women experienced higher mortality in major CS phenotypes while receiving less advanced mechanical circulatory support. These findings support early recognition, rapid phenotype classification, and sex-conscious but non-delayed escalation strategies for women with CS.

**Clinical Perspective:** *What Is New?:*

- In this national cohort of cardiogenic shock hospitalizations, sex-based differences in mortality were not uniform but varied substantially by shock phenotype.
- Women had higher mortality in major phenotypes, particularly acute myocardial infarction-related and heart failure-related cardiogenic shock.
- Women were less likely to receive mechanical circulatory support despite higher mortality in several clinically important phenotypes.

*What Are the Clinical Implications?:*

- Cardiogenic shock care should move beyond a uniform approach and incorporate early phenotype classification, structured risk assessment, and timely escalation pathways.
- For women, differences in presentation, comorbidity burden, vascular anatomy, bleeding risk, and procedural complications should inform procedural planning and risk mitigation, but should not delay escalation when advanced support is clinically indicated.
- Standardized shock-team activation, early hemodynamic assessment, bleeding-avoidance strategies, ultrasound-guided access, and alternative access planning may help reduce sex-based disparities in cardiogenic shock care.

## Introduction

Cardiogenic shock (CS) is a heterogeneous clinical syndrome characterized by low cardiac output and inadequate tissue perfusion. The Shock Academic Research Consortium (SHARC) defines CS in clinical practice as a cardiac disorder resulting in clinical and biochemical evidence of sustained tissue hypoperfusion, whereas clinical trials frequently use hemodynamic thresholds such as systolic blood pressure <90 mm Hg for more than 30 minutes or need for vasopressors in conjunction with signs of hypoperfusion [1].

The etiologies of CS include acute myocardial infarction (AMI), acute decompensated heart failure, arrhythmias, myocarditis, valvular heart disease, post-cardiotomy shock, and mixed or non-myocardial causes. The Society for Cardiovascular Angiography and Interventions (SCAI) staging system classifies shock severity from stage A (at risk) to stage E (extremis), providing a common framework for risk stratification, clinical communication, and comparison across studies [2].

Despite advances in pharmacologic support, early revascularization, invasive hemodynamic assessment, shock-team models, and temporary mechanical circulatory support (MCS), CS remains associated with high short- and long-term mortality [3-6]. Sex-based differences in cardiovascular disease presentation, treatment, and outcomes are well recognized, and several studies have reported that women with CS are older, have different comorbidity profiles, receive invasive therapies less frequently, and may experience worse short-term outcomes than men [7-10]. However, prior studies have often focused on AMI-related CS or treated CS as a single syndrome.

Because the mechanisms, treatment priorities, and risks of escalation differ across CS phenotypes, sex-based disparities may also differ by underlying phenotype. Defining these phenotype-specific differences is clinically important because it may inform early recognition, shock-team activation, invasive hemodynamic assessment, device selection, and procedural risk mitigation. We therefore used a nationally representative cohort to evaluate sex differences in mortality and treatment utilization across clinically relevant CS phenotypes.

## Methods

### Data Source

We performed a retrospective cohort study using the National Inpatient Sample (NIS), Healthcare Cost and Utilization Project, for 2019 through 2023. The NIS is a nationally representative all-payer database of United States hospital discharges and uses a stratified, cluster sampling design with discharge-level weights to generate national estimates. All analyses incorporated sampling weights, strata, and cluster variables according to HCUP recommendations. Because the NIS contains deidentified discharge-level data, this study was exempt from institutional review board review.

### Study Population and Phenotype Classification

Adult hospitalizations (age >=18 years) with CS were identified using ICD-10-CM code R57.0. CS hospitalizations were categorized into clinically relevant phenotypes using predefined ICD-10-CM codes: AMI (I21-I22), heart failure (I50), arrhythmia-related shock (I47.2, I49.0), myocarditis/Takotsubo (I40-I41, I51.81), valvular disease (I34-I39), and other etiologies. Cardiac arrest (I46) was included as a covariate rather than a defining phenotype. Phenotype definitions and coding hierarchy are provided in the Supplemental Methods and Supplemental Table 1.

### Exposure and Outcomes

The primary exposure was recorded sex (female versus male). The primary outcome was in-hospital mortality. Secondary outcomes included use of MCS, including extracorporeal membrane oxygenation (ECMO), intra-aortic balloon pump (IABP), Impella, composite IABP/Impella, and any MCS, as well as mechanical ventilation. Outcomes were evaluated overall and stratified by CS phenotype.

### Covariates

Covariates were selected a priori based on clinical relevance and prior literature and included age, diabetes mellitus, chronic kidney disease, coronary artery disease, chronic obstructive pulmonary disease, obesity, cirrhosis, acute kidney injury, cardiac arrest, PCI, CABG, mechanical ventilation, and MCS modalities. Mechanical ventilation was considered a marker of illness severity but may also represent a treatment-related variable influenced by practice patterns; therefore, sensitivity analyses were performed with and without adjustment for mechanical ventilation.

### Statistical Analysis

Survey-weighted analyses were used as the primary analytic approach. Categorical variables were compared using Rao-Scott chi-square tests, and continuous variables were compared using survey-weighted linear regression. Standardized mean differences (SMDs) were used to assess baseline differences, with SMD >0.10 considered clinically meaningful imbalance. Associations between sex and outcomes were evaluated using survey-weighted logistic regression models in the overall cohort and within CS phenotypes, with results reported as odds ratios (ORs) and 95% confidence intervals (CIs).

To further address confounding, we performed a propensity score-matched sensitivity analysis. A propensity score for female sex was estimated using multivariable logistic regression including demographics, comorbidities, CS phenotype, treatment variables, and illness severity markers. Patients were matched 1:1 using nearest-neighbor matching without replacement. Balance was assessed using SMDs, with SMD <0.10 indicating adequate balance. In the matched cohort, outcomes were compared using paired analyses and reported as absolute risk differences and risk ratios (RRs). All tests were two-sided, with statistical significance defined as p<0.05. Analyses were performed using software capable of complex survey estimation.

## Results

### Study Population

A total of 254,691 weighted hospitalizations with CS were identified. Of these, 158,747 (62.3%) occurred in men and 95,896 (37.7%) occurred in women. CS was categorized into six phenotypes: AMI-related, heart failure-related, arrhythmia-related, myocarditis/Takotsubo, valvular, and other etiologies. Heart failure-related CS was the most common phenotype (46.2%), followed by AMI-related CS (27.7%).

### Baseline Characteristics

Baseline characteristics differed significantly by sex (Table 1). Women were older than men (66.8 +/- 17.0 versus 64.6 +/- 15.8 years; SMD, 0.131; p<0.001). Men had higher prevalence of coronary artery disease (56.8% versus 44.7%; SMD, 0.244), acute kidney injury (66.4% versus 61.1%; SMD, 0.111), and chronic kidney disease (40.0% versus 35.7%; SMD, 0.089). Women had higher prevalence of obesity and chronic obstructive pulmonary disease. Clinically meaningful imbalances were observed for coronary artery disease, IABP/Impella use, CABG, age, acute kidney injury, and phenotype distribution (Table 2).

**Table 1.**
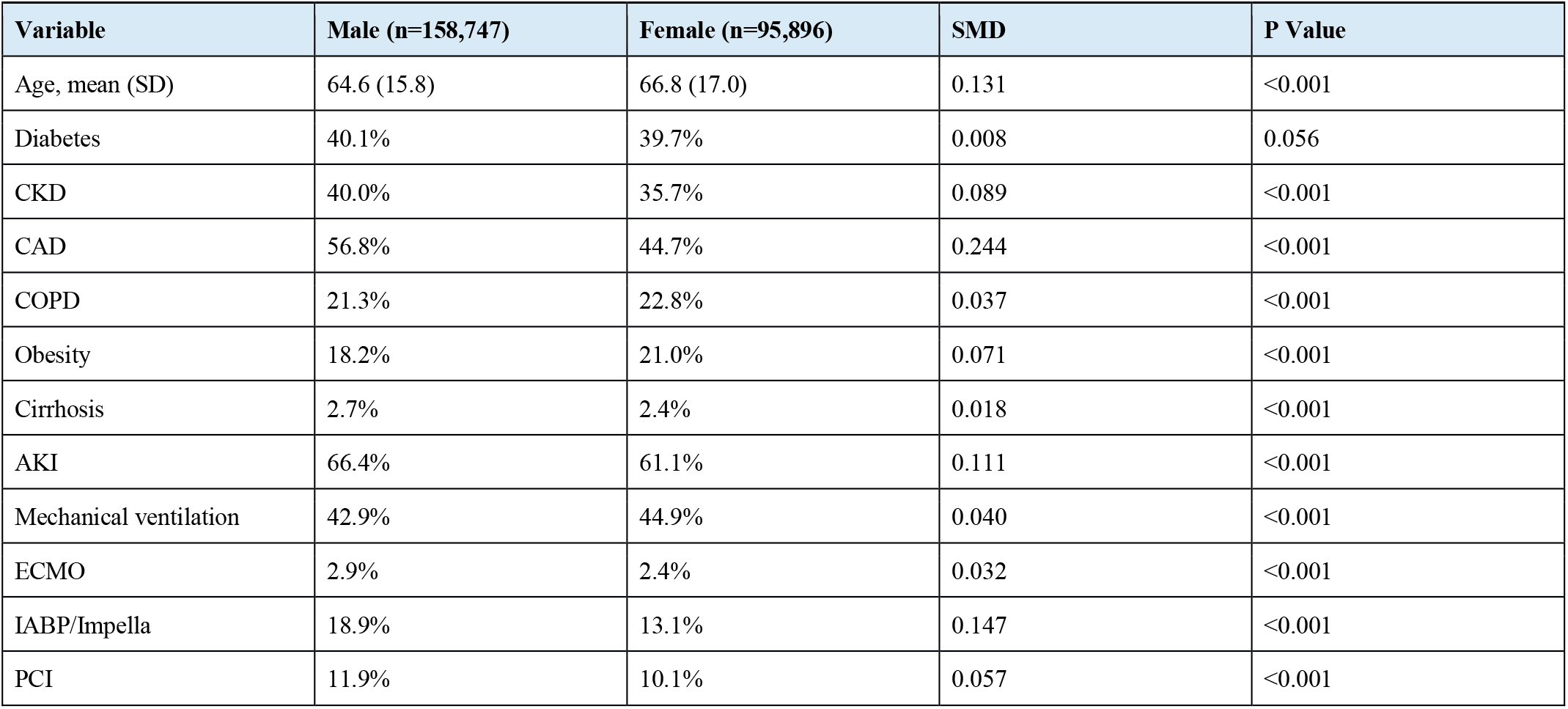
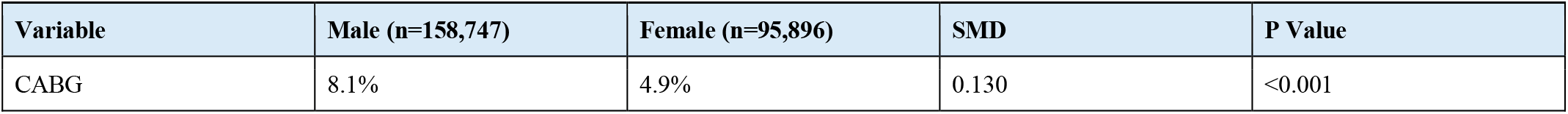
Baseline Characteristics by Sex.

**Table 2.** Variables With Meaningful Imbalance Before Matching.

| Variable | SMD |
| --- | --- |
| CAD | 0.244 |
| IABP/Impella | 0.147 |
| CABG | 0.130 |
| Age | 0.131 |
| AKI | 0.111 |
| Phenotype distribution | 0.108 |

### Treatment Utilization

Significant sex-based differences were observed in invasive therapies and MCS utilization. Women were less likely than men to receive ECMO (2.4% versus 2.9%; OR, 0.82; 95% CI, 0.77-0.87), IABP (12.7% versus 18.3%; OR, 0.65; 95% CI, 0.63-0.67), Impella (5.3% versus 8.4%; OR, 0.61; 95% CI, 0.58-0.63), IABP/Impella (13.1% versus 18.9%; OR, 0.65; 95% CI, 0.63-0.66), or any MCS (14.6% versus 20.4%; OR, 0.67; 95% CI, 0.65-0.68). Women were more likely to receive mechanical ventilation (44.9% versus 42.9%; OR, 1.08; 95% CI, 1.06-1.10).

### Survey-Weighted In-Hospital Mortality

In survey-weighted analyses, female sex was associated with higher in-hospital mortality in several CS phenotypes (Table 3). Compared with men, women had higher mortality in AMI-related CS (36.1% versus 31.3%; OR, 1.24; 95% CI, 1.19-1.28), arrhythmia-related CS (37.3% versus 31.6%; OR, 1.28; 95% CI, 1.20-1.38), and heart failure-related CS (30.5% versus 25.8%; OR, 1.27; 95% CI, 1.23-1.30). A smaller difference was observed in other CS phenotypes (OR, 1.05; 95% CI, 1.01-1.10), whereas no significant differences were observed in valvular or myocarditis/Takotsubo-related CS.

**Table 3.** Survey-Weighted In-Hospital Mortality by Phenotype.

| Phenotype | Male | Female | OR (Female vs Male) | P Value |
| --- | --- | --- | --- | --- |
| AMI | 31.3% (30.8-31.8) | 36.1% (35.3-36.8) | 1.24 (1.19-1.28) | <0.001 |
| Arrhythmia | 31.6% (30.7-32.6) | 37.3% (35.9-38.7) | 1.28 (1.20-1.38) | <0.001 |
| Heart failure | 25.8% (25.3-26.3) | 30.5% (30.0-31.1) | 1.27 (1.23-1.30) | <0.001 |
| Myocarditis/Takotsubo | 31.6% (23.2-40.0) | 38.3% (30.9-45.8) | 1.35 (0.82-2.23) | 0.235 |
| Valvular | 17.5% (13.9-21.1) | 18.5% (14.5-22.5) | 1.07 (0.75-1.53) | 0.701 |
| Other | 41.9% (41.1-42.8) | 43.1% (42.2-44.0) | 1.05 (1.01-1.10) | 0.024 |

**Table 4.** Mechanical Circulatory Support and Ventilation by Sex.

| Outcome | Male | Female | OR (Female vs Male) | P Value |
| --- | --- | --- | --- | --- |
| ECMO | 2.9% (2.7-3.0) | 2.4% (2.2-2.5) | 0.82 (0.77-0.87) | <0.001 |
| IABP | 18.3% (18.0-18.6) | 12.7% (12.4-13.0) | 0.65 (0.63-0.67) | <0.001 |
| Impella | 8.4% (8.2-8.6) | 5.3% (5.1-5.5) | 0.61 (0.58-0.63) | <0.001 |
| IABP/Impella | 18.9% (18.5-19.2) | 13.1% (12.8-13.4) | 0.65 (0.63-0.66) | <0.001 |
| Mechanical ventilation | 42.9% (42.4-43.4) | 44.9% (44.4-45.4) | 1.08 (1.06-1.10) | <0.001 |
| Any MCS | 20.4% (20.0-20.8) | 14.6% (14.3-15.0) | 0.67 (0.65-0.68) | <0.001 |

**Table 5.** Unadjusted Mortality Differences by Sex.

| Phenotype | Male Mortality | Female Mortality | Absolute Difference |
| --- | --- | --- | --- |
| AMI | 33.1% | 37.3% | +4.2% female |
| Arrhythmia | 39.4% | 40.4% | +1.0% female |
| Heart failure | 24.8% | 29.9% | +5.1% female |
| Myocarditis/Takotsubo | 20.1% | 21.4% | +1.3% female |
| Other | 44.7% | 44.7% | 0% |
| Valvular | 21.8% | 27.9% | +6.1% female |

### Propensity-Matched Analyses

After propensity score matching, baseline characteristics were well balanced between groups (all SMD <0.10). Mortality differences were attenuated but persisted in selected phenotypes. Women had higher mortality in valvular CS (absolute difference, +4.9%; RR, 1.21), heart failure-related CS (+3.0%; RR, 1.11), and AMI-related CS (+1.4%; RR, 1.04). In contrast, women had lower mortality after matching in arrhythmia-related CS (−2.1%; RR, 0.95) and other CS phenotypes (−2.3%; RR, 0.95). No significant difference was observed in myocarditis/Takotsubo-related CS. These findings suggest that baseline differences account for a substantial portion, but not all, of the observed sex differences in mortality.

## Discussion

### Principal Findings

In this nationally representative cohort of adults hospitalized with CS, sex differences in mortality and treatment utilization varied meaningfully by shock phenotype. Four principal findings emerged. First, women had higher in-hospital mortality in several major CS phenotypes, particularly AMI-related and heart failure-related CS. Second, women were less likely to receive MCS, including ECMO, IABP, Impella, and any MCS, despite higher mortality in key phenotypes. Third, women were more likely to receive mechanical ventilation, which may reflect greater illness severity, differences in clinical presentation, or variation in treatment pathways. Fourth, after propensity score matching, mortality differences were attenuated but persisted in selected phenotypes, supporting the importance of both baseline differences and potential differences in care processes.

### Phenotype-Specific Sex Differences in Cardiogenic Shock

Our findings support the concept that CS should not be treated as a single clinical entity when evaluating sex-based disparities. Prior literature has shown that women with CS often present at older age, have different comorbidity profiles, and experience higher short-term mortality than men [7-11]. A recent systematic review and epidemiologic meta-analysis reported higher unadjusted mortality among women and persistent excess risk after adjustment, while also demonstrating lower use of MCS among women [11]. Our analysis extends this work by showing that sex-based mortality differences are not uniform across CS phenotypes.

The phenotype-specific nature of our findings is clinically important. In AMI-related CS, women may experience delayed recognition, lower rates of early revascularization, more frequent non-ST elevation presentations, and higher rates of mechanical complications, all of which may contribute to adverse outcomes [7,10,13,14]. In heart failure-related CS, sex differences may reflect distinct patterns of chronic heart failure, pulmonary hypertension, renal dysfunction, frailty, and de novo etiologies such as myocarditis or stress cardiomyopathy. Contemporary registries have similarly emphasized that the characteristics and prognostic factors associated with CS differ between women and men [12,13].

### Treatment Utilization and Escalation of Care

The lower use of MCS among women in our cohort is consistent with prior national and registry data [11,12,15]. This finding should be interpreted carefully. Lower MCS use may reflect older age, comorbidity burden, smaller vascular caliber, bleeding risk, frailty, or clinical contraindications. However, it may also reflect delayed recognition, risk aversion, or variation in escalation practices. Because the NIS does not provide hemodynamics, SCAI stage, vasoactive medication dose, or timing of device placement, we cannot determine whether lower MCS use represented appropriate selection or potentially modifiable undertreatment.

Importantly, women-specific procedural risks should guide planning rather than delay escalation. Smaller access vessels, bleeding risk, vascular complications, renal dysfunction, and frailty are clinically relevant and should inform device choice, access strategy, and multidisciplinary planning. These considerations should prompt proactive mitigation strategies, including early vascular assessment, ultrasound-guided access, micropuncture techniques, careful sheath-to-femoral-artery assessment, closure planning, alternative access when appropriate, and early involvement of interventional cardiology, advanced heart failure, critical care, vascular surgery, and cardiothoracic surgery teams. They should not lead to therapeutic inertia when advanced support is otherwise indicated.

### Clinical Implications for Shock Systems of Care

These findings have practical implications for CS systems of care. Early recognition of CS in women may be particularly important because women may present with atypical symptoms, delayed diagnosis, higher comorbidity burden, or more advanced shock physiology at the time of recognition. Structured shock pathways that incorporate early hemodynamic assessment, rapid phenotype classification, and predefined escalation criteria may reduce subjective variation in care and help ensure that women are considered for invasive therapies with the same urgency as men.

A phenotype-specific approach is essential. AMI-related CS requires rapid revascularization decisions and consideration of unloading or support strategies. Heart failure-related CS requires assessment of congestion, right ventricular involvement, pulmonary hypertension, candidacy for temporary MCS, and potential transition to durable advanced therapies. Arrhythmia-related CS requires prompt rhythm control and evaluation of reversible triggers. Valvular CS requires early structural heart and surgical evaluation. In each phenotype, sex-conscious care should mean earlier risk recognition and better procedural planning, not delayed escalation.

### Limitations

This study has several limitations. First, as with all administrative database analyses, CS identification, phenotype assignment, comorbidities, procedures, and complications depend on ICD-10 coding and may be subject to misclassification. Second, the NIS does not include granular hemodynamic data, SCAI shock stage, laboratory values, vasoactive medication dose, timing of shock onset, timing of revascularization or MCS, device duration, vascular access site, or post-discharge outcomes. Third, mechanical ventilation, MCS, PCI, and CABG may function as both markers of illness severity and mediators of outcome, creating potential overadjustment in some models. Fourth, we could not determine whether lower use of MCS in women reflected appropriate clinical selection, delayed recognition, procedural contraindications, patient preference, institutional resources, or implicit bias. Finally, residual confounding is likely despite survey-weighted multivariable modeling and propensity score matching.

## Conclusions

In this nationally representative cohort, sex differences in CS outcomes and treatment utilization varied by underlying phenotype. Women experienced higher mortality in several major phenotypes, particularly AMI-related and heart failure-related CS, and were less likely to receive MCS. These findings emphasize the need to move beyond a uniform approach to CS and toward early phenotype classification, structured risk assessment, and timely escalation pathways.

For women with CS, early recognition may be particularly important given differences in clinical presentation, comorbidity burden, vascular anatomy, and procedural risk. Women-specific considerations, including bleeding risk, vascular complications, smaller access vessels, renal dysfunction, and frailty, should inform procedural planning and device selection but should not delay escalation when advanced support is clinically indicated. Future studies should incorporate hemodynamic data, SCAI shock stage, treatment timing, device strategy, procedural complications, and longitudinal outcomes to determine whether standardized, phenotype-specific shock pathways can reduce sex-based disparities in CS care and survival.

**Figure 1.**
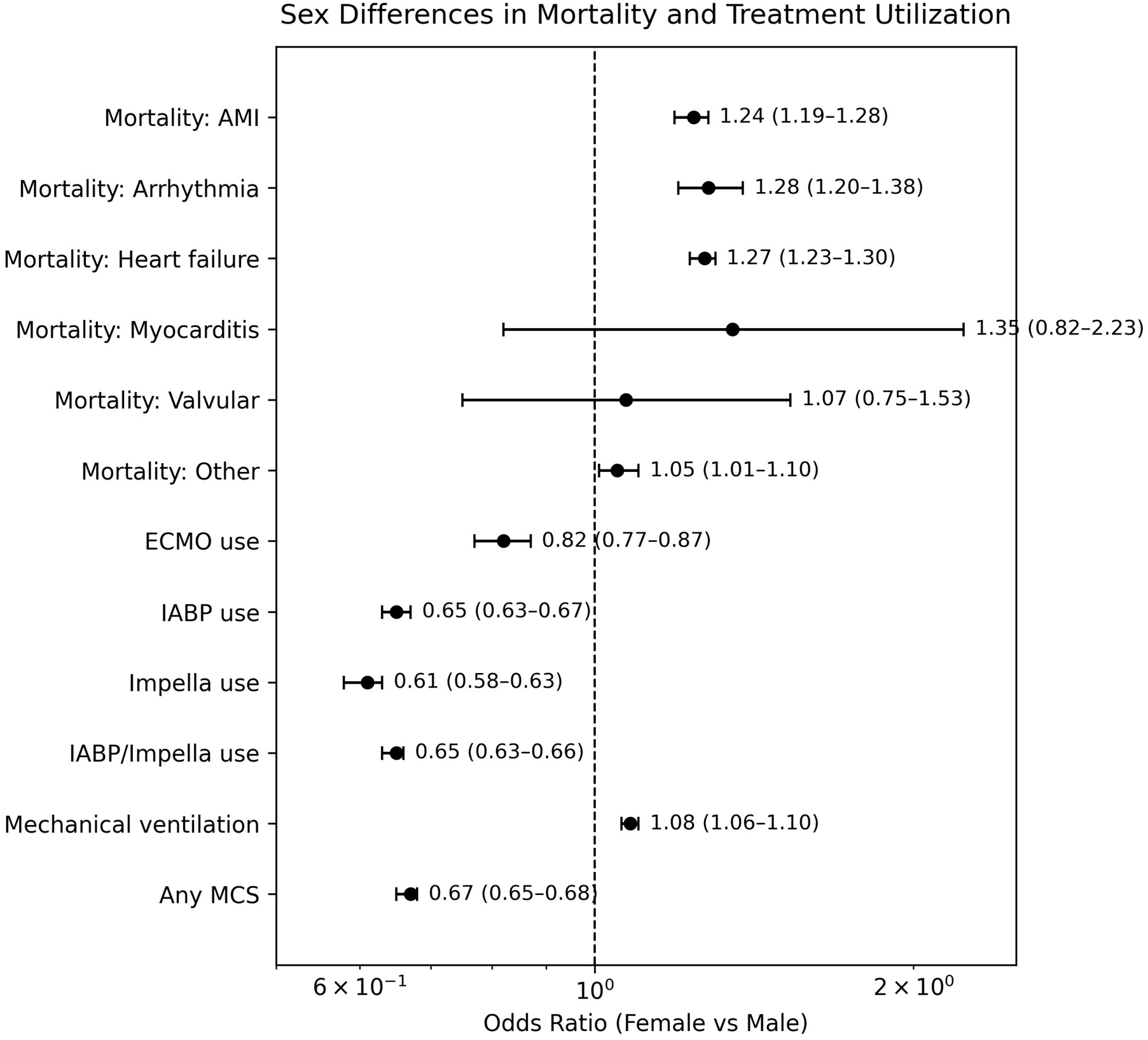
Forest plot of sex differences in mortality and treatment utilization. Odds ratios are shown for female versus male sex across phenotype-specific mortality outcomes and treatment utilization outcomes. Values greater than 1 indicate higher odds in women; values less than 1 indicate lower odds in women.

**Figure 2.**
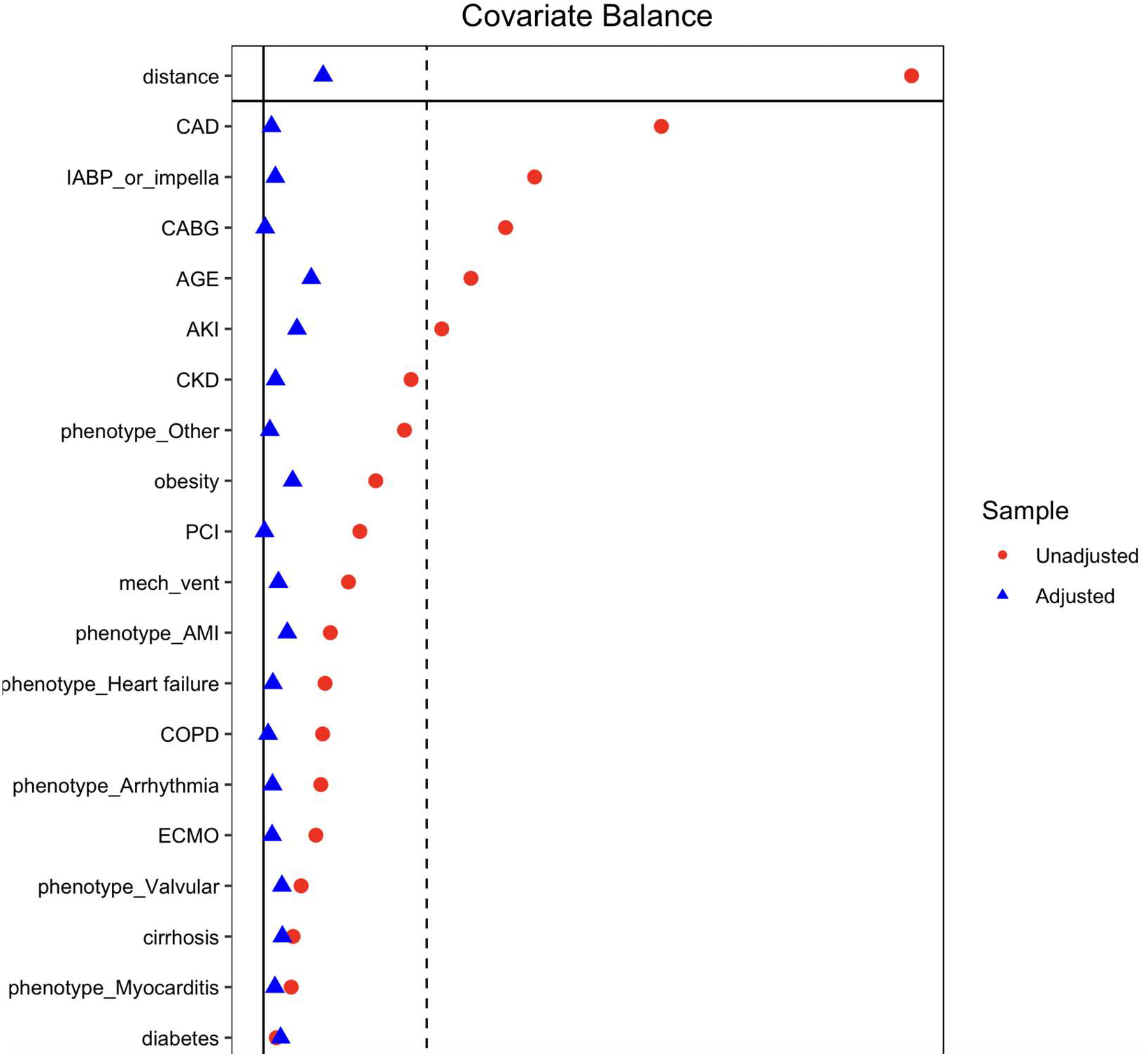
Covariate balance before and after propensity score matching. Standardized mean differences are shown before and after matching; values below 0.10 indicate adequate balance.

**Figure 3.**
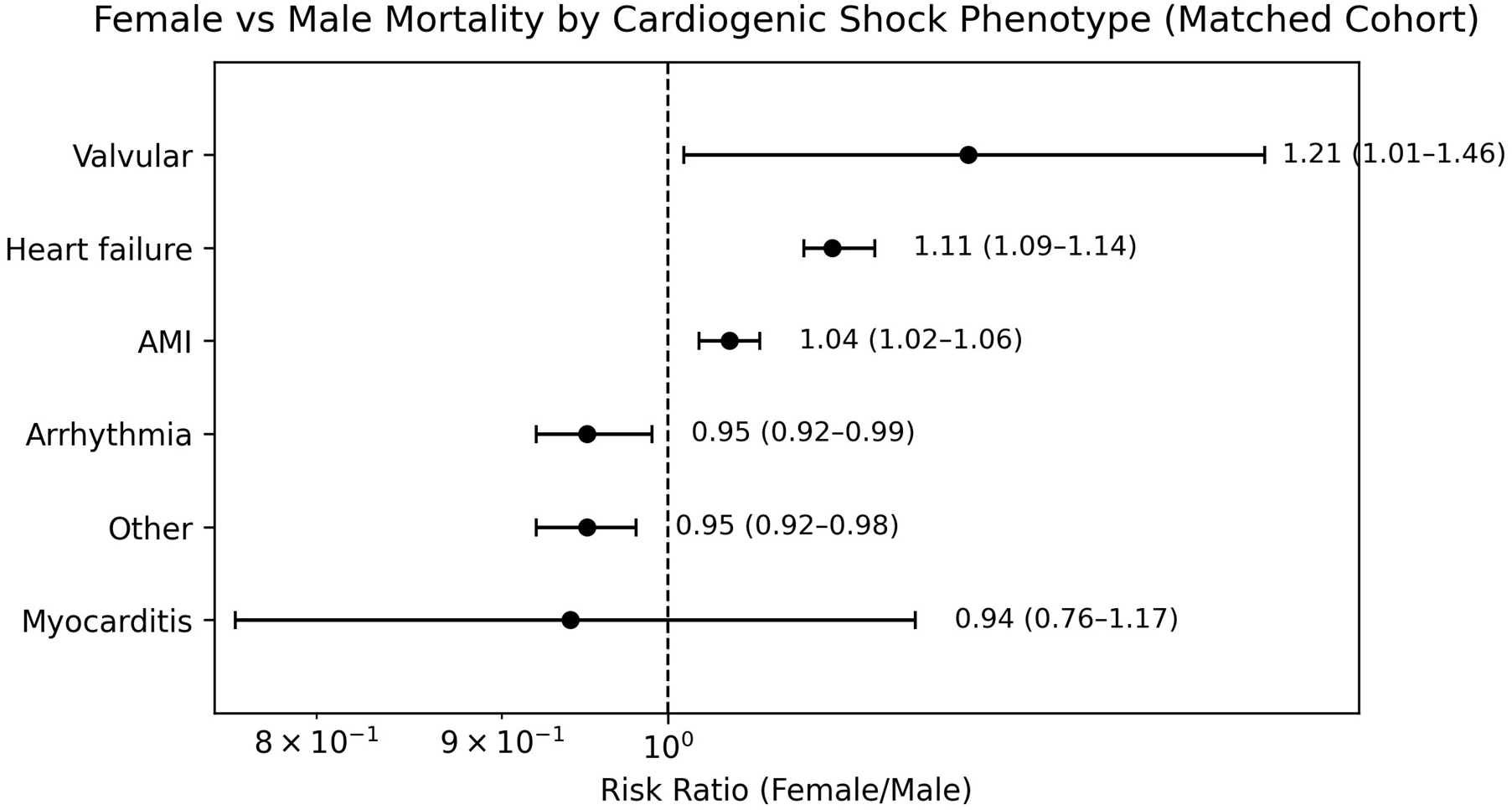
Propensity-matched mortality by cardiogenic shock phenotype. Risk ratios compare female versus male mortality after matching; values greater than 1 indicate higher mortality risk in women.

## Data Availability

Data is accessible to the public (NIS database) codes used are disclosed in the article

https://www.google.com/url?sa=t&source=web&rct=j&opi=89978449&url= https://hcup-us.ahrq.gov/nisoverview.jsp&ved=2ahUKEwjRlOn_78qUAxX9liYFHUctGpcQFnoECBkQAQ&usg=AOvVaw32negMDlaLa5UO4vpSpNSM

